# A Bi-Directional Mendelian Randomization Study on Five Major Psychiatric Disorders, Inflammatory Regulators, Brain Plasticity, and Related Traits

**DOI:** 10.1101/2025.10.08.25337604

**Authors:** Yuanxin Zhong, Yiming Qin, Zipeng Liu, Larry Baum, Thuan-Quoc Thach, Pak C. Sham

## Abstract

**Introduction:** Growing evidence links psychiatric disorders to immune system dysfunction, atypical brain development, and psychosocial traits such as intelligence and childhood maltreatment. However, the causal relationships between psychiatric disorders and these potential risk factors remain controversial.

**Objectives:** To better elucidate the intertwined pathways underlying five major psychiatric disorders and potential risk factors by employing genetic instruments to evaluate possible causal relationships.

**Methods:** We conducted bidirectional two-sample Mendelian randomization (MR) analyses by leveraging summary statistics from recent GWASs with large sample sizes. Causal relationships were estimated between five major psychiatric disorders (ADHD, ASD, BD, MDD, and SCZ) and various factors, including cytokines, longitudinal brain changes, childhood maltreatment, antisocial behavior, educational attainment, and intelligence, which were used as both exposures and outcomes (average N > 310k). LHC-MR, a novel MR method which controls for correlated horizontal pleiotropy and simultaneously estimate bi-directional causal effects for trait pairs, was utilized to discover potential causalities. MRCI, another advanced MR method for estimating reciprocal causal effects, and standard MR methods were employed to replicate the identified significant causalities. Moreover, potential mediating pathways were investigated.

**Results:** Causal relationships of 300 trait pairs were examined by LHC-MR, 41 of which achieved significance after multiple testing (p < 1.67e-4). After replication, LHC-MR, MRCI, and standard MR methods agreed on four positive causal effects, including attention-deficit/hyperactivity disorder (ADHD) on the chemokine RANTES and RANTES on major depression (MDD). Mediation analysis further supported that RANTES contributed to the association between ADHD and MDD. Additionally, both schizophrenia and MDD positively increased the risk of childhood maltreatment, perhaps due to premorbid traits which trigger bullying or poor parenting associated with genetic predisposition to these disorders.

**Conclusions:** The current study contributes to a better understanding of the intertwined causal network between psychiatric disorders and various risk factors, which may help develop public health prevention and intervention.

## Introduction

Psychiatric disorders pose a significant health burden on society, underscoring the urgent need for a deeper understanding of their pathogenic mechanisms. While the exact causes of psychiatric disorders remain largely unknown, SNP-based heritability estimates for disorders such as major depression (MDD) [1], attention-deficit/hyperactivity disorder (ADHD) [2], autism spectrum disorder (ASD) [3], bipolar disorder (BD) [4], and schizophrenia (SCZ) [5] range between 11% and 24%, highlighting the contribution of genetic components to these conditions. Moreover, previous research has revealed varying degrees of genetic overlap among pairs of these disorders [6], supported by observed patterns of shared associations with measures of immune response and structural/functional brain changes.

Mounting evidence indicates an intricate interplay between the immune system, as usually indexed by various inflammatory marker levels, and psychiatric disorders. Observational studies, including epidemiological and clinical studies, have reported multiple significant correlations [7-10]. However, the wide variability of the results has hampered definitive conclusions [10-13]. For example, an elevated level of interleukin-6 (IL-6) in ADHD patients compared to healthy individuals was found in one meta-analysis [10] but not another [13]. Similarly, while inflammatory markers have been extensively studied in MDD, and there is general agreement on the correlation of MDD with IL-6 and C-reactive protein (CRP), there is a lack of clarity with regard to other markers such as tumor necrosis factor-α (TNFα) and interleukin-1β (IL-1β), with substantial heterogeneity in correlation estimates across studies [11]. Furthermore, correlation does not imply causation, and there is as yet little consensus on the causal relationships between immune dysfunction and psychiatric disorders. Nevertheless, there is evidence that immune activation from prenatal or early postnatal infections contributes to increased SCZ risk [14, 15], and the higher risk of autoimmune diseases in patients with psychotic disorders suggests that the susceptibility or presence of psychiatric disorder may lead to immune dysfunction [16].

Neuroimaging studies have found changes in brain structure and function across multiple brain regions among individuals with psychiatric disorders. For example, studies have shown alterations in the regional brain volumes and cortical thickness in individuals with SCZ, BD, and MDD [17, 18]. Functional neuroimaging studies have also revealed changes in brain activity and connectivity patterns in psychiatric disorders [19, 20]. However, since most neuroimaging studies on psychiatric disorders are cross-sectional, the causality between brain changes and psychiatric disorder manifestation is difficult to establish. Additionally, immune cells and their products (e.g., cytokines) make essential contributions to brain development, modulating the proliferation, differentiation, and survival of neuronal cells, and synaptic growth and pruning [21, 22]. The potential mediation role of brain changes in the causal relationships between immune dysfunction and psychiatric disorders remains unclear.

In recent years, Mendelian Randomization (MR) has emerged as a powerful approach to examine the causality between two traits (exposure and outcome) when a randomized controlled trial (RCT) is not feasible [23]. MR analysis utilizes genetic variants as instrumental variables (IVs) to estimate the causal effect of the exposure on the outcome of interest [24] to overcome the problem of unmeasured confounding in traditional epidemiological studies. The availability of GWAS summary statistics on multiple psychiatric disorders, immune markers and brain imaging-derived phenotypes (IDPs) provides an excellent opportunity to apply MR to study causal relationships between them. In a systematic review, Iakunchykova, Leonardsen and Wang (25) identified 15 MR studies on immune dysfunction and psychiatric disorders published in the past 10 years. Among inflammatory markers, CRP and IL-6 have received the most attentions as potential causal factors of MDD and SCZ [26-28]. Moreover, a study reported causal effects of genetically predicted levels of IL-6 on grey matter volume and cortical thickness in the middle temporal gyrus and superior frontal region[29]. Another study provided evidence of causal effects of nine IDPs on risk of SCZ, BD and anorexia nervosa (AN) [30]. Relatively fewer studies have examined other markers [27, 31, 32], but these studies have nevertheless provided some interesting findings, including a putative causal effect of basic fibroblast growth factor (FGF-Basic) on AN, and BD on IL-9 [31].

However, for the causal inferences made by MR to be valid, three strong assumptions must be satisfied. When the exposure variable is itself a complex trait, multiple genetic instruments can be selected to serve as IVs, giving multiple estimates that can be combined by inverse variance weighting (IVW) [33]. However, as Iakunchykova, Leonardsen and Wang (25) pointed out, the highly polygenic nature of both inflammatory markers and psychiatric disorders means that some selected instruments may separately influence both exposure and outcome (horizontal pleiotropy) and violate MR assumptions. Refinements of standard methods have been proposed to increase the robustness to invalid instruments, e.g., MR Egger [34] and weighted median/mode [35]. However, these methods may not adequately protect against invalid causal inferences when the number of invalid instruments due to horizontal pleiotropy is large, or when the pleiotropic effects on exposure and outcome are correlated [36]. Among the 15 studies reviewed by Iakunchykova, Leonardsen and Wang (25), most did not adequately address the issue of correlated horizontal pleiotropy, making their bi-directional causal estimations by employing two separate analyses biased and problematic. Moreover, the availability of massive GWAS resources has led to the exponential explosion of MR papers containing many seemingly implausible causalities [37]. Owing to this extensive usage and the tendency of MR methodologies to violate model assumptions, there is a real potential for false positive causal inferences in such studies which could lead to ineffective or even harmful public health policies. The successful application of MR may require it to be placed in a triangulation of evidence framework, involving the integration of various causal inference methods and multiple study designs and data source to tackle the same issue[37].

In this study, we aim to untangle the causal relationships between psychiatric disorders and cytokines, brain plasticity, and psychosocial traits. To address the recognized limitations and enhance the robustness of the causal estimations, we leveraged summary statistics from recent large-scale GWASs, adopted a two-stage study design and employed an advanced MR technique for discovery, Latent Heritable Confounder MR (LHC-MR), an advanced method which incorporates a latent variable to allow for potential correlated horizontal pleiotropy while simultaneously estimating bi-directional causal effects. We applied the Bonferroni correction to adjust the significance level for multiple testing. The significant results were further replicated using an in-house developed method, Mixture model Reciprocal Causation Inference (MRCI) which also performs simultaneous reciprocal causality estimation, and standard MR methods. Furthermore, we applied the two-step MR model to test the potential mediation effects, which allowed us to explore the causal relationships in a more comprehensive manner.

## Materials and Methods

### Data sources

We obtained the latest GWAS summary statistics for five major psychiatric disorders from the Psychiatric Genomics Consortium (PGC) (https://pgc.unc.edu/for-researchers/download-results/) for SCZ (76,755 cases and 243,649 controls) [5], MDD (246,363 cases and 561,190 controls) [1], BD (41,917 cases and 371,549 controls) [4], ADHD (38,691 cases and 186,843 controls) [2], and ASD (18,381 cases and 27,969 controls) [3] (Supplementary Table 1). Information regarding genotyping, imputation, quality control, and genetic association analysis was described in the primary research papers for each disorder.

For immune dysfunction, we utilized cytokine measures indexing chronic inflammation. The latest GWAS summary statistics for 41 inflammatory cytokines (n = 8,293) were incorporated into the analyses [38]. This cross-sectional study examined data from three cohorts, including the Cardiovascular Risk in Young Finns Study (YFS, n = 1,980) (https://youngfinnsstudy.utu.fi/) and the FINRISK 1997 (n = 4,608) and 2002 cohorts (n = 1,705) (www.thl.fi/finriski). Semi-fasting EDTA plasma, semi-fasting heparin plasma, and fasting serum were used for the cytokine quantification in FINRISK 1993, FINRISK 2002, and YFS, respectively. After quality control and merging the datasets, 41 cytokine measures were available in YFS and FINRISK 2002, and 17 out of 41 measures were also available in FINRISK 1997 (Supplementary Table 2). The association models were generated from fitting the first ten genetic principal components, age, and sex as covariates. Further details regarding the data can be found in the primary paper [38].

The GWAS summary statistics of changes in brain morphology across the lifespan from ENIGMA-Plasticity Working Group were obtained (n = 15,640) (Supplementary Table 1). Brain measures were extracted from longitudinal magnetic resonance imaging (MRI) data. Annual rates of changes for 15 brain structures (ml or cm^2 per year), including eight global brain measures and seven subcortical structures, were computed by subtracting baseline brain measures from follow-up measures and dividing by the number of years of follow-up duration. Overall head size was not corrected. Further details about included cohorts, quality control of MRI data, meta-analysis and mega-regression have been described in the primary study [39].

Additionally, we included four psychosocial traits in the analysis. The GWAS summary statistics for childhood maltreatment (n = 185,414), antisocial behavior (n = 85,359), educational attainment (n = 1,131,881), and intelligence (n = 269,867) were obtained from four large-scale studies [40-43] (Supplementary Table 1). Detailed information can be found in the primary research papers for each trait. GWAS summary statistics were obtained from different consortia or studies, which reduced the risk of bias caused by sample overlap.

### Ethics statement

In this study, only publicly accessible GWAS data were utilized. The original GWAS study had obtained ethical approval and consent for participation.

### Statistical analysis

#### MR analysis: discovery stage

We first used LHC-MR to simultaneously estimate bi-directional causal effects between five major psychiatric disorders and 41 cytokines, brain plasticity for 15 brain structures measured by annual rates of change, and four psychosocial traits (childhood maltreatment, antisocial behavior, educational attainment, and intelligence). Briefly, LHC-MR incorporates a latent variable as the potential heritable confounder which affects exposure and outcome in a structural equation model (SEM) to control correlated horizontal pleiotropic effect. Maximum likelihood estimates for bidirectional causal effects, confounder effects, direct heritabilities, and others are obtained by optimized likelihood function. The standard errors of these estimates are calculated using the block jackknife approach implemented in LHC-MR [44]. Additionally, LHC-MR overcomes some limitations such as potential sample overlap and insufficient utilization of genome-wide markers.

We used the default parameter settings in LHC-MR in the estimation. We conducted a total of 300 tests and applied the Bonferroni correction to account for multiple testing. A p < 0.05/300 = 1.67e-4 was considered statistically significant. However, if exposure of any trait pair having less than 3 valid IVs (i.e., variants satisfying the MR assumptions), the corresponding result would be considered as a lack of robustness unless one of the other MR methods agreed with it.

#### MR analysis: replication stage

For detected trait pairs with significant causal links, we utilized MRCI [45] and standard MR methods to validate the causal directions and effects.

Similar to LHC-MR, MRCI also estimates reciprocal causalities simultaneously. But, instead of selecting valid IVs, MRCI makes use of GWAS summary statistics of all available SNPs of the trait pair and groups them into four categories (trait-specific, pleiotropic, and null SNPs) which explicitly models pleiotropy. Then it constructs a composite likelihood function accounting for LD among SNPs based on external reference LD information. For MRCI, the simulation results showed that the estimations are robust to pleiotropic causal variants [45].

For the standard MR method, we prioritized IVW in the estimations. We also explored weighted median and MR Egger regression for causal estimation. If exposure of any trait pair had a single valid IV, the Wald method was applied. We extracted the valid IVs included in MR tests for each trait pair by LHC-MR, input them as the selected IVs when using standard MR methods, and compared the causal estimates with those from LHC-MR and MRCI. To check the robustness of the results, we performed a series of sensitivity analyses, including Cochran’s Q test to detect heterogeneity among selected IVs, and computed MR Egger regression intercept to estimate horizontal pleiotropy. We generated scatter plots, forest plots and funnel plots, and conducted leave-one-out analyses, and computed approximate F-statistics by equation of {3^2^/se^2^ [46] to detect outliers among included IVs. In the presence of heterogeneity or/and horizontal pleiotropy among IVs, estimations using the weighted median and MR Egger were prioritized rather than IVW. Additionally, Chen, Yao, Cai, Fu, Li and Wu (31) utilized the same datasets of psychiatric disorders and cytokines as the current study and performed standard MR analyses following the regular pipeline. We also checked the result consistency between Chen, Yao, Cai, Fu, Li and Wu (31) and the current study.

#### Two-step MR analysis

We adopted two-step MR to assess whether the exposure-outcome causal link is mediated by other risk factors. Two-step MR analysis consists of two analyses. The first analysis is to detect the causality between exposure and the potential mediator, and the second is to detect the causal relationship between the potential mediator and the outcome of interest. To investigate the mediation effect of cytokines and brain plasticity, we conducted two-step MR analysis when the causal effects of the same psychiatric disorder (as an exposure) on cytokines/brain plasticity and another non-overlap phenotype were detected, such as significant causal effects of SCZ (exposure) on IL-6 (potential mediator) and SCZ on intelligence (outcome), or significant causal effects of MDD (exposure) on annual rate of change in hippocampus (potential mediator) and MDD on childhood maltreatment (outcome).

Before examining the effect of the potential mediator on the outcome of interest (step two), we first excluded all IVs used in the first step, which were the SNPs used in MR to determine the causal effect of psychiatric disorders on the potential mediator, and SNPs in high LD with the included IVs (r^2^ > 0.8 based on the 1000 Genomes Project European reference panel) from the GWAS summary statistics of the potential mediator. Then, we input the re-generated GWAS summary statistics of the potential mediator and GWAS summary statistics of the outcome of interest into LHC-MR and standard MR methods to estimate the causal relationship between them. If a significant causality between the potential mediator and the outcome of interest was present, we used the Product method to estimate the beta coefficient of the indirect effect [47] and the Delta method to estimate standard error (SE) and 95% confidence intervals (CIs) of the indirect effect [48].

The overall pipeline of the current study is summarized in Figure 1.

**Figure 1.**
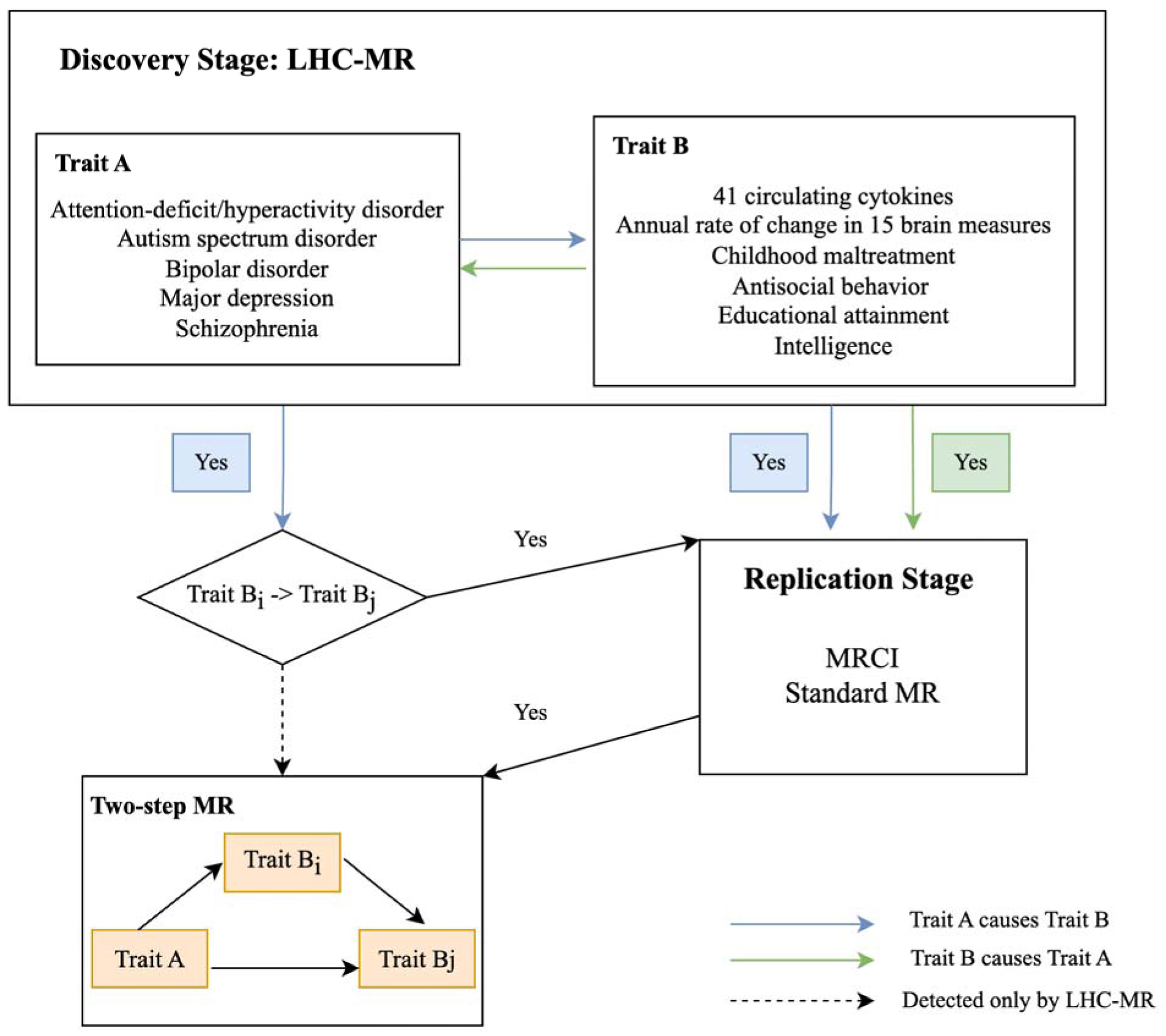
The overall pipeline of current study.

## Results

### Discovery stage: significant causality detected by LHC-MR

We tested for causal relationships between 300 trait pairs, and we found significant causal relationships for 41 trait pairs, including 11 for ADHD, 2 for ASD, 7 for BD, 14 for MDD, and 7 for SCZ (Figure 2 and Supplementary Table 3-5).

**Figure 2.**
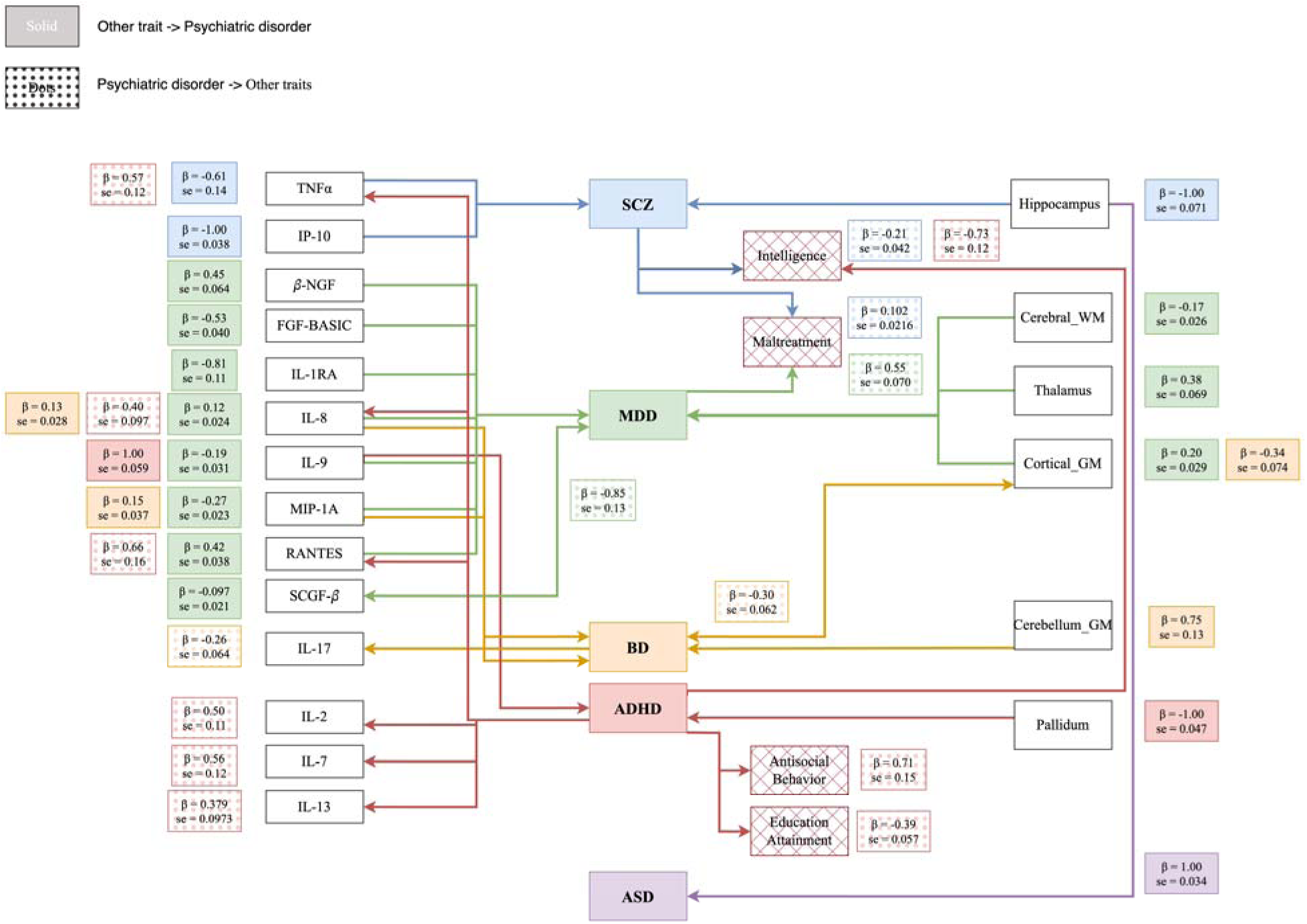
Significant causal relationship for 41 trait pairs identified by LHC-MR. SCZ, schizophrenia, its causalities were colored in blue; MDD, major depression, its causalities were colored in green; BD, bipolar disorder, its causalities were colored in orange; ADHD, attention-deficit/hyperactivity disorder, its causalities were colored in red; ASD, autism spectrum disorder, its causalities were colored in purple; Cerebellum GM, rate of change in cerebellar grey matter; Cortical GM, rate of change in cortical grey matter; Pallidum, rate of change in pallidum; Cerebral WM, rate of change in cerebral white matter; Thalamus, rate of change in thalamus; Hippocampus, rate of change in hippocampus.

### Replication stage

#### Consistent results across different MR methods

Among significant causal relationships for 41 trait pairs detected by LHC-MR, MRCI identified consistent causal estimations for causality of six trait pairs, and standard MR methods agreed with causation for 11 trait pairs (Supplementary Figures 1-3 and Supplementary Table 6). LHC-MR, MRCI, and standard MR methods agreed on four causalities, including the causal effects of (1) ADHD on chemokine RANTES; (2) chemokine RANTES on MDD; (3) SCZ on childhood maltreatment; and (4) MDD on childhood maltreatment.

In sensitivity analyses, among these four casualties, we observed a significant heterogeneity among IVs for RANTES when inferring a causal effect on MDD, SCZ on childhood maltreatment, and MDD on childhood maltreatment (all p < 0.05). We found no evidence of horizontal pleiotropy for IVs (all p > 0.05, see Supplementary Material). So, for standard MR methods, we prioritized estimations from the weighted median (Table 1). The results of F-statistics for selected IVs for all trait pairs could be found in the Supplementary Material (Supplementary Table 8-47).

**Table 1.** Consistent results across LHC-MR, MRCI, and standard MR.

| Causality | LHC-MR |  | MR-CI |  | Method | Standard MR |  |
| --- | --- | --- | --- | --- | --- | --- | --- |
| | $\beta$ (SE) | P | $\beta$ (SE) | P | | $\beta$ (SE) | P |
| RANTES -> MDD | 0.42(0.038) | 1.57E-27 | 0.26(0.032) | 3.83E-16 | Weighted median | 0.010(5.0E-3) | 3.53E-02 |
| ADHD -> RANTES | 0.66(0.16) | 3.95E-05 | 0.066(0.018) | 2.05E-04 |  | 0.79(0.36) | 2.86E-02 |
| SCZ -> Childhood maltreatment | 0.10(0.022) | 2.31E-06 | 0.13(0.058) | 2.71E-02 |  | 0.071(0.012) | 1.66E-09 |
| MDD -> Childhood maltreatment | 0.55(0.069) | 1.91E-15 | 0.69(0.076) | 8.98E-20 |  | 0.41(0.064) | 1.35E-10 |

Note: MDD, Major depression; ADHD, attention-deficit/hyperactivity disorder; SCZ, schizophrenia; RANTES, chemokine RANTES.

#### Partially consistent results across different MR methods

There were nine causations identified by two MR methods. Two of these causations were identified by LHC-MR and MRCI analyses, whereas standard MR methods agreed with LHC-MR on the causal estimations for causality of seven trait pairs (Table 2 and Supplementary Figure 3). Sensitivity analysis result for each trait pair can be found in the Supplementary Material.

**Table 2.** Significant causalities partially consistent across MR methods.

| Causality | MR Methods | $\beta$ (SE) | P |
| --- | --- | --- | --- |
| BD -> IL-17 | LHC-MR | -0.26(0.064) | 5.66E-05 |
|  | MRCI | -0.075(0.0090) | 3.32E-16 |
| BD -> Cortical GM | LHC-MR | -0.30(0.062) | 9.48E-07 |
|  | MRCI | -0.096(0.02) | 1.66E-06 |
| IP10 -> SCZ | LHC-MR | -1.00(0.038) | 1.26E-150 |
|  | Weighted median | -0.035(0.014) | 1.55E-02 |
| MDD -> SCGF-β | LHC-MR | -0.85(0.13) | 1.32E-11 |
|  | IVW | -0.85(0.28) | 2.73E-03 |
| SCZ -> Intelligence | LHC-MR | -0.21(0.042) | 8.33E-07 |
|  | Weighted median | -0.057(0.012) | 1.38E-06 |
| ADHD -> Intelligence | LHC-MR | -0.73(0.12) | 4.02E-10 |
|  | Weighted median | -0.33(0.055) | 1.25E-07 |
| ADHD -> Antisocial behavior | LHC-MR | 0.71(0.15) | 3.71E-06 |
|  | IVW | 0.54(0.057) | 1.78E-11 |
| ADHD -> Educational attainment | LHC-MR | -0.39(0.057) | 1.78E-11 |
|  | Weighted median | -0.17(0.029) | 3.59E-09 |
| Hippocampus -> ASD | LHC-MR | 1.00(0.034) | 2.07E-193 |
|  | IVW | 0.045(0.022) | 4.0E-2 |

Note: ADHD, attention-deficit/hyperactivity disorder; ASD, autism spectrum disorder; BD, bipolar disorder; MDD, major depression; SCZ, schizophrenia; Cortical GM, rate of change in cortical grey matter; Hippocampus, rate of change in hippocampus.

Additionally, Chen, Yao, Cai, Fu, Li and Wu (31) followed the standard MR pipeline and identified the consistent causal effects with LHC-MR in the current study for ADHD on IL-7 [IVW: {3 = 0.234, se = 0.103, p = 0.023; LHC-MR: {3 = 0.555, se = 0.123, p = 6.65e-6], and ADHD on TNFα [IVW: {3 = 0.209, se = 0.102, p = 0.041; LHC-MR: {3 = 0.574, se = 0.121, p = 2.21e-6].

#### Discordance between different MR methods

Seven trait pairs showed a disagreement between estimations computed by LHC-MR and by other MR methods in the current study (Table 3). Additionally, Chen, Yao, Cai, Fu, Li and Wu (31) found negative causal effects of MDD on {3-NGF (IVW: {3 = -0.297, se = 0.144, p = 0.040) and 1L-1RA on MDD (IVW: {3 = -0.017, se = 6.6e-3, p = 0.01), and a positive causal effect of FGFBasic on MDD (IVW: {3 = 0.031, se = 0.012, p = 0.009).

**Table 3.**
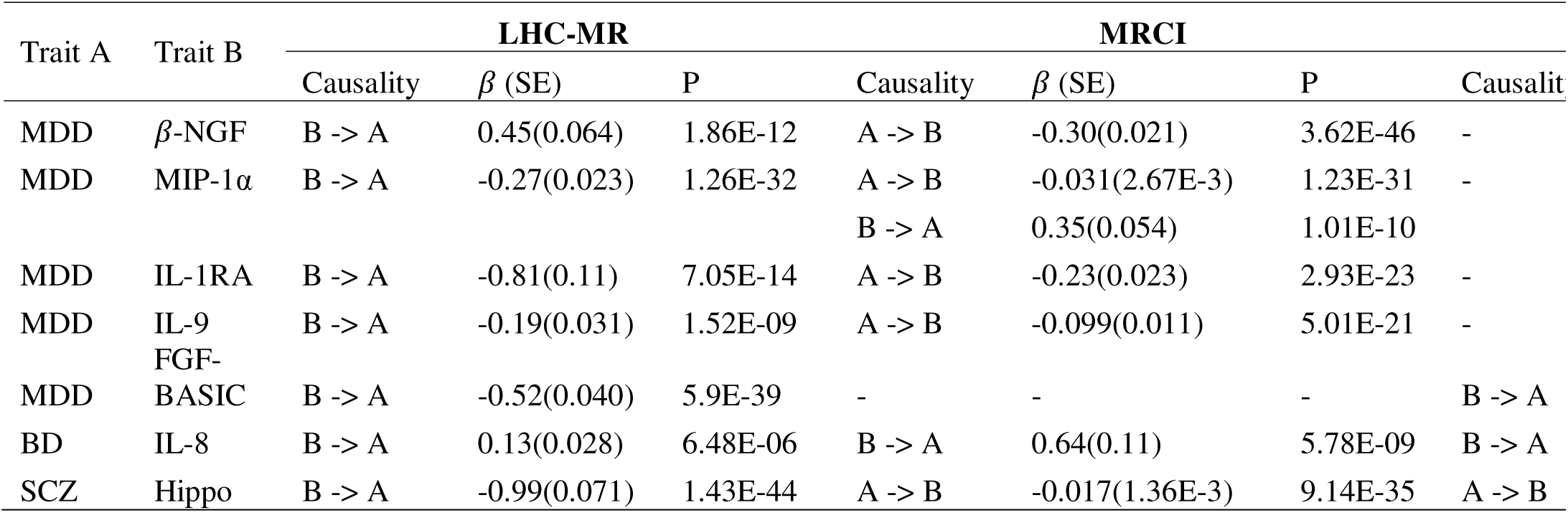
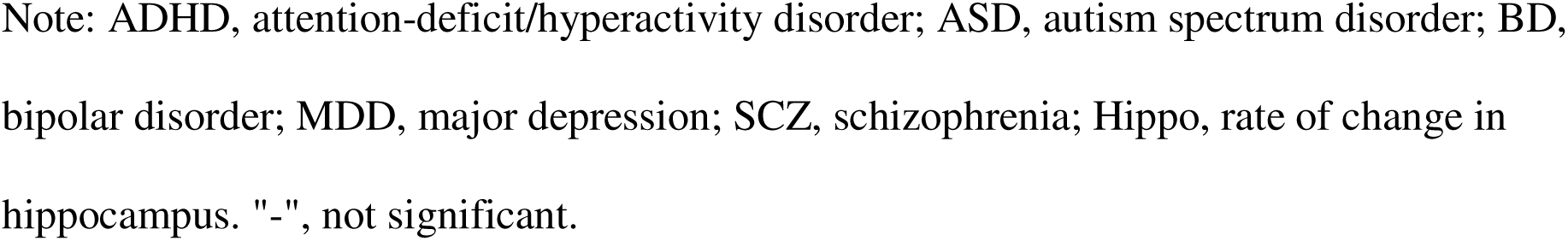
Inconsistent bi-directional causal estimations across different MR methods.

| Trait A | Trait B | LHC-MR |  |  | MRCI |  |  |  |
| --- | --- | --- | --- | --- | --- | --- | --- | --- |
| | | Causality | $\beta$ (SE) | P | Causality | $\beta$ (SE) | P | Causality |
| MDD | β-NGF | B -> A | 0.45(0.064) | 1.86E-12 | A -> B | -0.30(0.021) | 3.62E-46 | - |
| MDD | MIP-1α | B -> A | -0.27(0.023) | 1.26E-32 | A -> B | -0.031(2.67E-3) | 1.23E-31 | - |
|  |  |  |  |  | B -> A | 0.35(0.054) | 1.01E-10 | - |
| MDD | IL-1RA | B -> A | -0.81(0.11) | 7.05E-14 | A -> B | -0.23(0.023) | 2.93E-23 | - |
| MDD | IL-9 | B -> A | -0.19(0.031) | 1.52E-09 | A -> B | -0.099(0.011) | 5.01E-21 | - |
| MDD | FGF-BASIC | B -> A | -0.52(0.040) | 5.9E-39 | - | - | - | B -> A |
| BD | IL-8 | B -> A | 0.13(0.028) | 6.48E-06 | B -> A | 0.64(0.11) | 5.78E-09 | B -> A |
| SCZ | Hippo | B -> A | -0.99(0.071) | 1.43E-44 | A -> B | -0.017(1.36E-3) | 9.14E-35 | A -> B |

Note: ADHD, attention-deficit/hyperactivity disorder; ASD, autism spectrum disorder; BD, bipolar disorder; MDD, major depression; SCZ, schizophrenia; Hippo, rate of change in hippocampus. "-", not significant.

### Two-step MR analysis

A significant positive causal effect of ADHD on MDD was captured by LHC-MR (p = 6.49e-29) and standard MR methods (all p < 0.05). We identified that the level of RANTES significantly partially mediated the causal relationship between ADHD and MDD (Figure 3). Additionally, since the causal effects of ADHD on RANTES, ADHD on TNFα, ADHD on intelligence, ADHD on antisocial behavior, ADHD on educational attainment, MDD on SCGF-{3, MDD on childhood maltreatment, BD on IL-17, and BD on rate of change for cortical grey matter were supported by more than one MR method, we examined the causal effects of (1) RANTES and TNFα on intelligence, antisocial behavior and educational attainment; (2) SCGF-{3 on childhood maltreatment; and (3) IL-17 on rate of change for cortical grey matter, to explore the potential mediation effect of (1) RANTES and TNFα on causalities of ADHD; (2) SCGF-{3 on causality of MDD; and (3) IL-17 on causality of BD (Supplementary Table 7). We identified potential causal effects of TNFα on educational attainment and on intelligence by LHC-MR, suggesting the mediation effect of TNFα on the causalities between ADHD and intelligence, as well as between ADHD and educational attainment. However, the standard MR methods did not produce consistent causality results, suggesting that these significant results were not robust (Supplementary Table 7).

**Figure 3.**
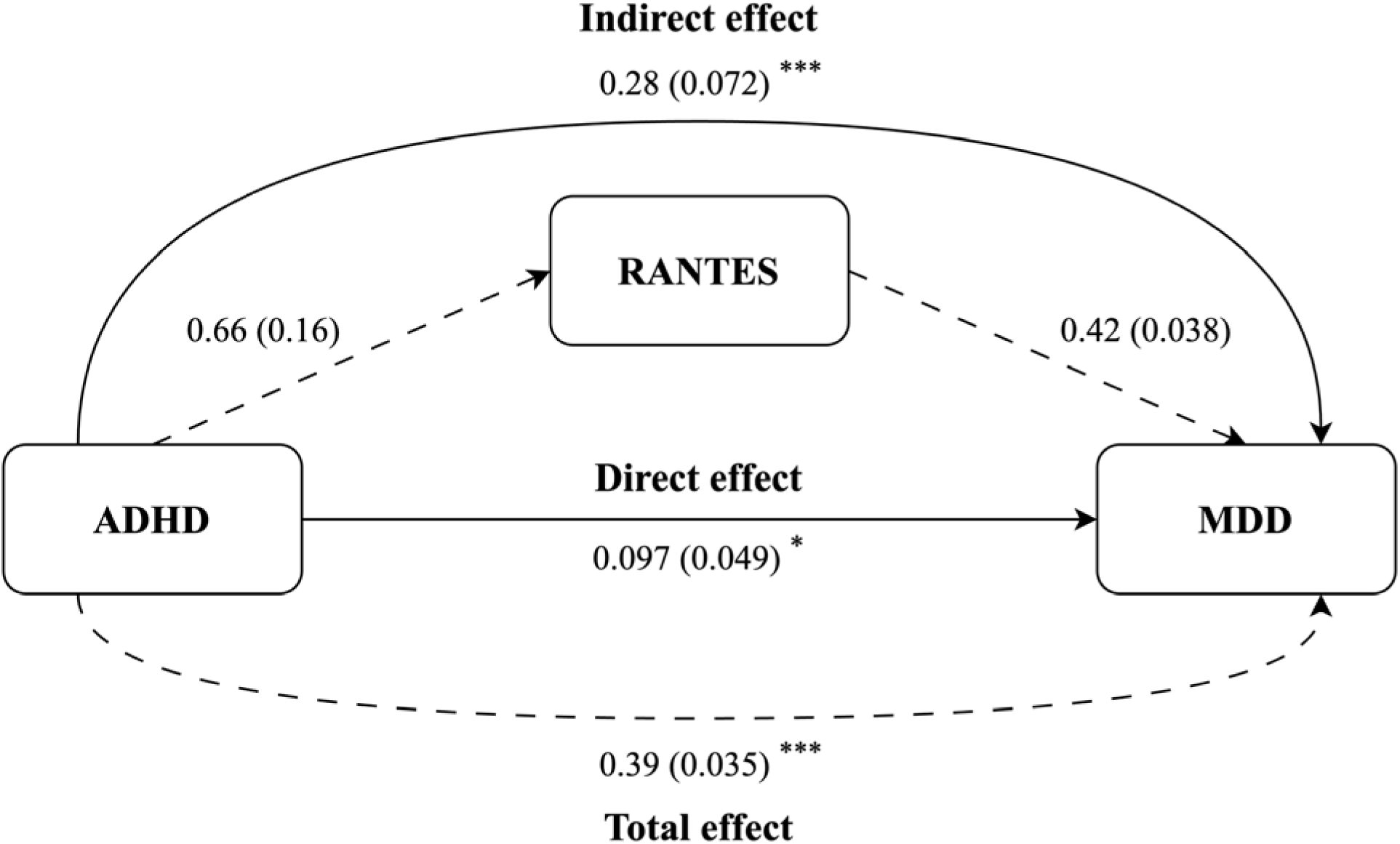
Results of two-step MR analyses for ADHD, MDD and RANTES, showing the direct and indirect pathways within the mediation model. The arrow direction indicates the causal direction. The values were formatted as (SE). *, p < 0.05; ***, p < 0.001.

## Discussion

In the current study, we leveraged summary statistics from recent GWASs. These examined five major psychiatric disorders (SCZ, MDD, BD, ADHD, and ASD), 41 circulating cytokines, childhood maltreatment, antisocial behavior, educational attainment, intelligence, and rates of change for eight global brain measures and seven subcortical structures. To untangle the causal relationships of psychiatric disorders with other risk factors, we utilized standard MR methods, as well as two novel MR methods that can simultaneously estimate the reciprocal causation. Although discrepancies between the results across different MR methods were observed, we identified consistent estimates for four causalities: the causal effects of RANTES on MDD, ADHD on RANTES, SCZ on childhood maltreatment, and MDD on childhood maltreatment. We failed to find a robust mediation effect of cytokines on the causality between psychiatric disorders and other phenotypes, but the results suggested that TNFα potentially mediated the causal effect of ADHD on intelligence and educational attainment.

RANTES was named after a fictional psychiatric patient (from the 1986 movie "Man Facing Southeast"), which makes the associations of RANTES with psychiatric disorders coincidentally appropriate. RANTES, also known as chemokine ligand 5 (CCL5), is expressed and secreted by T cells upon activation. Acting as one of the important chemoattractants, RANTES recruits and activates certain types of inflammatory cells, particularly monocytes and T cells, in response to infection or tissue damage[49]. A key role in MDD for a protein with functions in the immune system is not surprising given the intimate involvement of immunity and MDD [7]. In the context of psychiatric disorders such as MDD and SCZ, studies observed altered expression of RANTES and suggested it may serve as a potential biomarker and target for treatment [50, 51]. Another study examined the correlation of MDD therapy response and cognitive function changes with alterations in mRNA levels of RANTES and one of its receptors, chemokine receptor 5 (CCR5). Patients who responded to treatment had lower levels of CCR5 and RANTES relative to non-responders, and an increased misperception of the emotion “anger” and a faster perception of the emotion “disgust” were linked to decreases in CCR5 and RANTES, respectively [52]. RANTES may block activation of GPR75, a G-protein coupled receptor, by 20-HETE (a metabolite of arachidonic acid) [53]. Disrupting GPR75 reduces body weight in humans and mice and inhibits exploration in mice [54]. Decreased interest in activities, such as eating or leaving the home, are symptoms of MDD [55]. These observational studies suggested the involvement of RANTES in the development of MDD, which was supported by the causal effect of RANTES on MDD in the current study.

Regarding ADHD, limited research has found a link between ADHD and RANTES [56, 57]. Antioxidants might act as an adjuvant ADHD therapy [58], consistent with a role for RANTES in ADHD. The detected causal effect of ADHD on RANTES may be explained by the progressive changes in ADHD over time [59]. Additionally, ADHD medication, allergic inflammation, and lifestyle factors, especially chronic stress, could also cause an increase of inflammatory cytokines in patients with ADHD [60, 61].

Epidemiological studies have reported robust associations of childhood maltreatment with the subsequent development of schizophrenia spectrum disorders and depression [62-64]. While this provides some support for childhood maltreatment to be a causal risk factor for these psychiatric disorders, the possible confounding from gene-environment correlation needs to be considered. Firstly, an increased predisposition to psychiatric disorders, which is genetically transmitted to children, may also increase the risk of maltreatment towards children [65, 66]. Thus, genetic associations with disease could be explained by confounding with genetic nurture[67], and familial environment associations with disease could be explained by genetic transmission. There is evidence for parental mental health problems (including SCZ and MDD) to have detrimental effects on parenting [65, 66], raising the possibility for genetic transmission to contribute to the association between childhood maltreatment and psychiatric disorder. Secondly, it is increasingly recognized that children are not passive receivers of parental care, but interact with their parents to alter their emotions and behavior. Thus, a child with increased predisposition to SCZ and MDD may exhibit traits that increase the risk of maltreatment by parents[68]. Such premorbid childhood traits could contribute to the association between childhood maltreatment and adult psychopathology. A recent review on MR[23] made the point that, when the putative exposure of a significant MR result is dichotomous (as is the case for SCZ and MDD), it is better to consider the genetic liability to the exposure (rather than the exposure itself) as being causal, to allow for the possibility that the true cause is some unmeasured variable with a strong genetic correlation to the disorder. From the “nature” perspective, individuals with a higher genetic risk of developing psychosis may manifest some behavior or manner that could trigger maltreatment in the pre-psychotic phase[68]. From the “genetic nurture” perspective, parents of psychotic patients may have a greater genetic susceptibility of developing psychiatric disorders and a higher probability of being diagnosed with them since offspring inherit genetic information from their parents[69]. Parents with psychotic diagnoses or exhibit premorbid traits could have a higher risk of parenting poorly, including all types of childhood maltreatment, leading to an increase in externalized behavior problems in offspring over time [66]. Studies on the causations between childhood maltreatment and psychiatric disorders have yielded inconsistent results, possibly due to the intertwined effects of nature and genetic nurture factors [67]. The evidence presented in our study hints at the causal direction and underscores the necessity for assessment and intervention of premorbid traits in both offspring and parents, which may lower the incidence of childhood maltreatment. However, further exploration is required to confirm these findings.

Consistent with a high genetic correlation between ADHD and MDD [6], the revealed mediation pathway between ADHD, RANTES and MDD suggests that RANTES may serve as a mediator contributing to the relationship between ADHD and MDD. Other causalities identified by two-step MR methods were supported by observational studies to a certain extent. For example, Izumi, Hino, Wada, Nagaoka, Kawamura, Mori, Sainouchi, Kakita, Kasai, Kunii and Yabe (70) observed a decreased expression level of IP-10 in patients with SCZ, which, to a degree, is consistent with the negative causal effect of IP-10 on SCZ in the current study. Secondly, a systematic review of longitudinal studies that show the association between greater loss of grey matter in frontal brain regions and mood episodes in BD patients as time progresses aligns with the negative causal effect of BD on rate of change for cortical grey matter [71]. Additionally, the links between SCZ and intelligence [72], ADHD and intelligence [73], ADHD and educational attainment [74], and between ADHD and antisocial behavior [75] have been much discussed. Some observational studies show discordant results [76, 77], which may be due to the differences in samples, methods and measures. However, the current study shows the potential causality inferred from genetic data, which is much less confounded by other factors, and may give direction to future studies.

The discrepancies between the results across different MR methods and the causality detected by a single MR method could be attributed to the limitations of each MR method. Standard MR methods are vulnerable to correlated horizontal pleiotropy, which violates the second assumption—independence. Such pleiotropic effects could cause severe bias to the estimation. Although LHC-MR incorporates a latent variable as the potential heritable confounder which affects exposure and outcome in a SEM to control for a correlated horizontal pleiotropic effect, its single confounder assumption may be broken by trait pairs with numerous confounders and uneven effect ratios, which might result in biased causal effect estimates. Furthermore, if the genetic basis of the target traits differs from a two-component Gaussian mixture of effect sizes, LHC-MR is not the best option [44]. Regarding MRCI, its assumption of bivariate normality of standardized marginal effects in the four SNP components may be violated when applied to real data, affecting the causal estimation. A second limitation of MRCI is that the calculation of robust sandwich standard errors, which is necessary for compositing likelihood for model fitting, could make the tests overly conservative and decrease their power. MRCI may not adequately handle the scenario where very strong pleiotropic effects and nearly no exposure-specific causal SNPs exist [45]. Additionally, summary statistics with relatively small sample sizes, such as for cytokines and brain imaging, may lead to biased causal estimates.

In conclusion, the current study contributes to a better understanding of the causal relationship between psychiatric disorders and cytokines, brain plasticity, and other traits including intelligence, childhood maltreatment, antisocial behavior, and educational attainment. Further work on the causal inter-relationships among ADHD, RANTES and MDD, and among childhood maltreatment, MDD and, SCZ, may help develop effective public health prevention and intervention strategies for psychiatric disorders.

## Data availability

All GWAS summary statistical data in this study are publicly available (listed in the Supplementary Table 1 & 2). All software packages used are publicly available.

